# Fatality assessment and variant risk monitoring for COVID-19 using three new hospital occupancy related metrics

**DOI:** 10.1101/2022.02.03.22270417

**Authors:** Ping-Wu Zhang, Steven H. Zhang, Wei-Feng Li, Casey J. Keuthan, Shuaizhang Li, Felipe Takaesu, Cynthia A. Berlinicke, Jun Wan, Jing Sun, Donald J. Zack

## Abstract

**Background:** Though case fatality rate (CFR) is widely used to reflect COVID-19 fatality risk, it’s use is limited by large temporal and spatial variation. Hospital mortality rate (HMR) is also used to assess the severity of COVID-19, but HMR data is not directly available except 35 states of USA. Alternative metrics are needed for COVID-19 severity and fatality assessment.

**Methods:** New metrics and their applications in fatality measurements and risk monitoring are proposed here. We also introduce a new mathematical model to estimate average hospital length of stay for death (*L*_*dead*_) and discharges (*L*_*dis*_). Multiple data sources were used for our analysis.

**Findings:** We propose three new metrics, hospital occupancy mortality rate (HOMR), ratio of total deaths to hospital occupancy (TDHOR) and ratio of hospital occupancy to cases (HOCR), for dynamic assessment of COVID-19 fatality risk. Estimated *L*_*dead*_ and *L*_*dis*_ for 501,079 COVID-19 hospitalizations in US 34 states between Aug 7, 2020 and Mar 1, 2021 were 14.0 and 18.2 days, respectively. We found that TDHOR values of 27 countries are less spatially and temporally variable and more capable of detecting changes in COVID-19 fatality risk. The dramatic changes in COVID-19 CFR observed in 27 countries during early stages of the pandemic were mostly caused by undiagnosed cases. Compared to the first week of November, 2021, the week mean HOCRs (mimics hospitalization-to-case ratio) for Omicron variant decreased 34.08% and 65.16% in the United Kingdom and USA respectively as of Jan 16, 2022.

**Interpretation:** These new and reliable measurements for COVID-19 that could be expanded as a general index to other fatal infectious diseases for disease fatality risk and variant-associated risk monitoring.

**Research in context:** *Evidence before this study:* We searched PubMed, medRxiv, and bioRxiv for peer-reviewed articles, preprints, and research reports on risk and health care evaluation for COVID-19 using the search terms “hospital occupancy mortality rate”, “ratio of total deaths to hospital occupancy”, “ratio of hospital occupancy to case” up to Jan 20, 2022. No similar concepts or studies were found. No similar mathematical models based on “hospital occupancy mortality rate” for the estimation of hospital length of stay for deaths and discharges have been identified to date.

*Added value of the study:* Our new metrics, HOMR and TDHOR, mimic HMR for COVID-19 fatality risk assessment but utilize readily available data for many US states and countries around the world. HOCR mimics hospitalization-to-case ratio for COVID-19. We also provide evidence that explains why COVID-19 CFR has such dramatic changes at the beginning of a COVID-19 outbreak. We have additionally provided new metrics for COVID-19 fatality risk dynamic monitoring including Omicron variant and showed that these metrics provided additional information.

*Implications of all the available evidence:* The results of this study, including average hospital length of stay for deaths and discharges for over 500,000 COVID-19 hospitalizations in the US, can aid county, state, and national leaders in making informed public health decisions related to the ongoing COVID-19 pandemic. This is the first study to provide quantitative evidence to address why CFR has a such a large variation at the beginning of the COVID-19 pandemic in most countries and will hopefully encourage more countries to release hospital occupancy data, which we show is both useful and easy information to collect. The new metrics introduced by our study are effective indicators for monitoring COVID-19 fatality risk, as well as potentially fatal COVID-19 variants, and could also be expanded to other fatal infectious diseases.

## Introduction

The case fatality rate (CFR), which represents the fraction of individuals with a particular disease who die from that disease, is one of the most commonly used metrics for assessing the severity of infectious disease outbreaks. CFR has been important in aiding county, state, and national leaders in making informed public health decisions related to the ongoing COVID-19 pandemic, as well as management of many other outbreaks such as those involving influenza, SARS-CoV-1 and MERS-CoV, diseases in which the CFR has ranged from as little as 0.12% to as high as 32.7% ^1-3^. CFR can have large temporal and spatial variation. For example, reported COVID-19 CFR values have varied from 0.048% (Singapore) to 10.16% (Mexico) as of Oct 20, 2020^4^. CFR can also vary highly even within an individual country or region at different stages of a disease outbreak^5^. A multitude of factors could potentially contribute to regional CFR incongruities, including patient access to health care, testing capacity, age, race, sampling, vaccination status, personal compliance to government guidance, and evolving SARS-CoV-2 variants^6-9^.

RNA viruses, which include SARS-CoV-2, are more rapidly mutating than DNA viruses. New viral variants raise widespread concern, especially when the mutations cause substantial changes in antigenicity, transmissibility, and virulence. SARS-CoV-2 variants of concern, including Alpha (B.1.1.7), Beta (B.1.351), Gamma (P.1), Delta (B.1.617.2), and the recently emerged highly infectious Omicron (B.1.1.529) variant, which have already spread around the world^10-15^.

The world’s COVID-19 experience has clearly demonstrated the reliance of informed public health decision making on having available easy, accurate, and reliable methods to rapidly monitor fatality and hospitalization risk of newly emerging variants and diseases. To address this need, and improve existing methods, we here propose hospital occupancy mortality rate (HOMR), ratio of total deaths to hospital occupancy (TDHOR) and ratio of hospital occupancy to cases (HOCR) as three, new, alternative and complementary measurements for COVID-19 fatality risk evaluation, early CFR variation analysis, and dynamic monitoring.

## Results

### Concepts of three new hospital occupancy related metrics and their relationships to CFR and HMR

Hospital mortality rate (HMR) is commonly used as an indicator of patient safety and quality of care in healthcare facilities, and it is also being used to assess the severity of the COVID-19 pandemic^16-17^. COVID-19 hospital deaths and cumulative hospitalization data are needed to calculate HMR; however, this information is not available for most countries, and is only available for 35 states in the US^18^. The US, China, Canada, Israel, Malaysia, Australia and approximately half of the European countries (29 countries total) have, on the other hand, released continuous daily hospital occupancy data^19^. Is there any way to link daily hospital occupancy to cumulative hospitalization? We found that the sum of daily hospital occupancy (HO) for a period and cumulative hospitalization can be bridged together by average length of stay for hospitalizations (*L*_*mean*,_ for both deaths and discharges). That means “sum of daily hospital occupancy” equals *L*_*mean*_ times cumulative hospitalizations for a specific period.

If HMR for COVID-19 is a ratio of hospital deaths to cumulative hospitalizations, what is a ratio of hospital deaths to the sum of daily hospital occupancy for a period? We propose this to be hospital occupancy mortality rate (HOMR), which is the daily HMR of the hospital stay period since HMR equals HOMR times *L*_*mean*_. HOMR is a new hospital occupancy related metric and may mimic HMR to a certain degree since it is directly related to HMR through *L*_*mean*_.

As noted above, CFR variation for COVID-19 can arise from variation in deaths, cases, or both. Some countries and USA states provide COVID-19 death data broken down into hospital deaths, long term facility deaths, and deaths at home. China and another 27 countries only release total deaths. To address this limitation in available data, we introduce a concept of ratio of total deaths to the sum of daily hospital occupancy (TDHOR) for a specific period. In contrast to hospitalization-to-case ratio (HCR, a ratio of cumulative hospitalizations to cases), the ratio of hospital occupancy to cases (HOCR) is an additional hospital occupancy metric derived in this study and HOCR can mimic HCR since HOCR equals HCR times *L*_*mean*_. Together, HOMR, TDHOR and HOCR provide three new hospital occupancy related metrics.

In regional population screening in mainland China, asymptomatic cases have been reported to represent 38.23% of total cases from Apr 18, 2020 to Jan 23, 2022 (Appendix Table 1). Such information is generally not available since in most parts of the world COVID-19 cases were found not from population screening but rather from diagnosed cases with symptoms, which equals total symptomatic cases minus undiagnosed cases. The CFR mentioned in the present study refers to symptomatic CFR if not otherwise specified^20^. The intrinsic relationships among CFR, HMR, and these three new metrics, are shown in formulas 1-2 below (Methods 1, formulas 3-13):

Formula 1: 1/HOMR

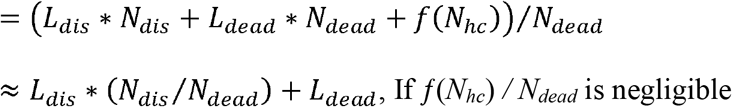

Formula 2: Case fatality rate (CFR) = total deaths /cases

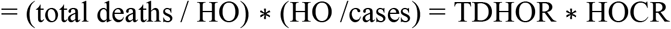

Notes for the formulas: 1) *L*_*mean*_, *L*_*dis*_ and *L*_*dead*_ are length of stay for hospitalizations (in-patients), discharges and deaths, respectively; 2) *N*_*dead*_, *N*_*dis*_ and *N*_*hc*_ are in-patient numbers for deaths, discharges and patients currently in hospital; 3) HO is hospital occupancy for a period (Methods 1, Formula 5); 4) The *f*(*N*_*hc*_) is the sum of period hospital stay for *N*_*hc*_. Usually *N*_*hc*_ < sum of recently new admitted patients for *n* days, *n* is close to *L*_*mean*_ and *f*(*N*_*hc*_) < *N*_*hc*_* *L*_*mean*_.

### Average hospital length of stay for COVID-19 deaths (*L*_*dead*_) and discharges (*L*_*dis*_) were estimated using HOMR in US

Length of stay in the hospital for deaths and discharges are useful fatality risk measurements for the COVID-19 pandemic because they reveal risk information for severe cases, which is missing in the CFR calculation. Several studies have addressed this at the beginning of the COVID-19 outbreak^21-22^. *L*_*dead*_ and *L*_*dis*_ can be estimated if we know HOMR and HMR for multiple days, since 1/HOMR≈ *L*_*dis*_ * (*N*_*dis*_/ *N*_*dead*_) + *L*_*dead*_ = *L*_*dis*_ * (1/HMR - 1) + *L*_*dead*_ (combination of formula 1 and 4).

There were 34 states within the US that we could use publicly available hospital deaths and cumulative hospitalization data for analysis, with 174,167 deaths and 501,079 hospitalizations within this period (Table 1). Interestingly, the regression plot for these states revealed different correlations at three different time periods (Fig.1A): Jun 26, 2020 to Aug 6, 2020 (42 days), Aug 7, 2020 to Nov 15, 2020 (101 days), and Nov 16, 2020 to Mar 1, 2021 (106 days). While the latter two time periods had nearly perfect linear correlation (r^2^ values of 0.97 and 0.99), for the first period and overall the correlations were limited (r^2^ values of 0.57 and 0.75, respectively). Estimated *L*_*dead*_ and *L*_*dis*_ for the latter two time periods are showed in Table 1 based on their intercepts of the Y axis and slope (Fig.1B-D). These data indicated that *L*_*dead*_ and *L*_*dis*_ were constant within these two periods, and the changes between them were subtle. At the time of Feb 15, 2021, only 4.24% of population was fully vaccinated against COVID-19 in the US, so our estimations should not likely be affected by vaccination rates^4^.

**Table 1.** Hospital occupancy data and estimation of average length of hospital stay in the US.

|  | Aug 7, 2020 - Nov 15, 2020 | Nov 16, 2020 - Mar, 1, 2021 | Combined |
| --- | --- | --- | --- |
| **Hospital occupancy** | 2,400,088 | 4,891,845 | 7,291,935 |
| **Total deaths** | 46,774 | 127,393 | 174,167 |
| **Hospital deaths** | 28,621 | 90,953 | 119,574 |
| **Hospitalizations** | 164,391 | 336,688 | 501,079 |
| **HOMR (%)** | 1·19 | 1·86 | 1·64 |
| **TDHOR (%)** | 1·95 | 2·6 | 2·39 |
| **HMR (%)** | 17·41 | 28·72 | 23·78 |
| **Length of stay in hospital for discharges (day)** | 14·3(95%CI:13·8-14·9) | 13·8(95%CI:13·6-14·0) | 14·0(95%CI:13·9-14·0) |
| **Length of stay in hospital for deaths (day)** | 20·3(95%CI:17·4-23·3) | 17·5(95%CI:16·6-18·5) | 18·2(95%CI:17·9-18·5) |
| **Correlation ($r^2$)** | 0.97 | 0.99 | NA* |
| **$L_{mean}$**<br>**(period hospital occupancy/**<br>**cumulative hospitalizations)** | 15·3(95%CI:15·2-15·3) | 14·5(95%CI: 14·4-14·5) | 14·9(95%CI:14·8-14·9) |

\*NA, Not Applicable.

Can these estimations can be supported by other data? *L*_*mean*_ is equal to hospital occupancy divided by cumulative hospitalizations and these are independent results. Our estimations for lengths of stay in the hospital for deaths (*L*_*dead*_) and discharges (*L*_*dis*_) matched the *L*_*mean*_ (14.9± 0.4, Table 1). Moreover, our estimated LOS for deaths and discharges were very close to what was previously reported by Verity et al., who found the mean duration from onset of symptoms to death (*T*_*od*_) and recovery (*T*_*or*_) for severe cases of COVID-19 in mainland China were 24.7 days and 17.8 days, respectively^21^. Similar studies estimated the mean time from onset to death (*T*_*od*_) was 20.0 days ^22^, and the average time from onset to hospitalization (*T*_*oh*_) was 7.0 days^23, 24^. Based on these estimations and formula *L*_*dead*_ =*T*_*od*_*-T*_*oh*_ and *L*_*dis=*_ *T*_*or*_*-T*_*oh*_, we calculated *L*_*dead*_ and *L*_*dis*_ and found that the Verity’s estimation *L*_*dead*_= 17.8-7.0=10.8 days and *L*_*dis*_= 24.7-7.0=17.7 days, while the Wu’s estimation was *L*_*d*_ =20.0-7.0=13 days. Our estimated *L*_*dead*_ was 14.0 days, which close to Wu’s *L*_*dead*_ estimation (13.0 days, based on Chinese data before Apr, 2020) and our estimated *L*_*dis*_ (18.2 days) was similar to Verity’s *L*_*dis*_ estimation (17.7 days).

We next wanted to know whether HOMR correlated with HMR in the US. To confirm this, we calculated *L*_*mean*_ for the combined 34 states and found that the *L*_*mean*_ is very steady (14.9 days, 95%CI:14.8-14.9) for a period of 7 months, spanning from Aug 7, 2020 to Mar 1, 2021 (Fig. 1E). Thus, HMR, which is a fatality risk index for hospitalizations, can be mimicked by HOMR in these states within a 7-month period (HMR = HOMR\**L*_*mean*_). The coefficient of determination was 0.943 (Fig. 1F). The COVID-19 mean HMR in this period (23.78%) was 2.98-fold that of the US CDC-estimated HMR for seasonal flu for the 2019-2020 period^2^.

**Fig. 1.**
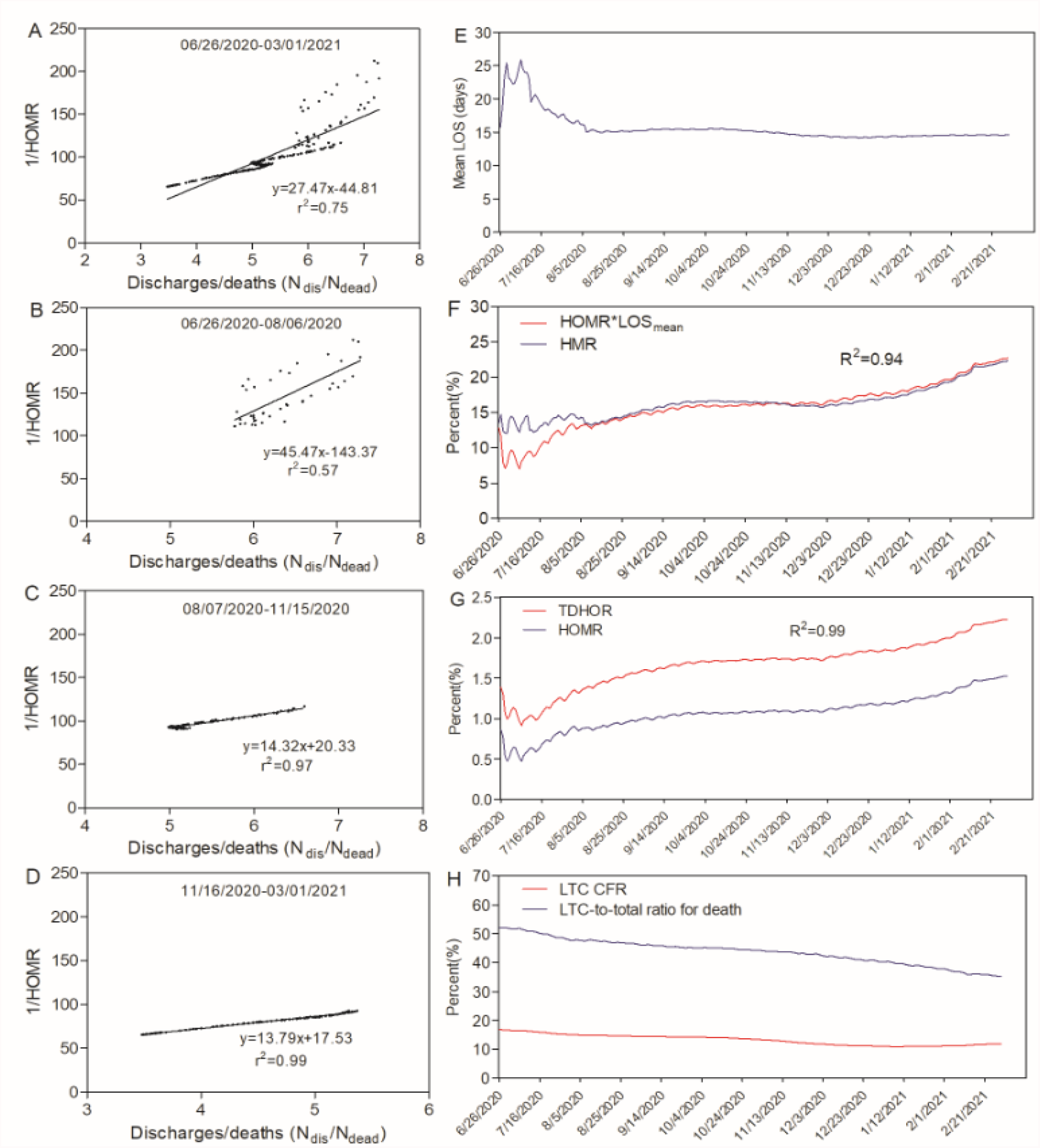
Estimations of average hospital length of stay for COVID-19 deaths and discharges and its correlation with HMR and TDHOR within the US. **A-D**, Scatterplots of COVID-19 data from 34 US states combined showing the relationship between 1/HOMR and *N*_*dis*_*/N*_*dead*_. Linear regression was used to estimate average length of hospital stay for COVID-19 deaths and discharges. Estimations were made for the COVID-19 infection time course of **A**, Jun 26, 2020 to Mar 1, 2021, as well as shorter time periods **B**, Jun 26, 2020 to Aug 6, 2020, **C**, Aug 7, 2020 to Nov 15, 2020, and **D**, Nov 15, 2020 to Mar 1, 2021. The linear regression equations and r^2^ values are included when applicable. **E**, *L*_*mean*_ calculations for the combined 34 states from Jun 26, 2020 to Mar 1, 2021. **F**, COVID-19 HMR and HOMR\**L*_*mean*_ for US states between Jun 26, 2020 and Mar 1, 2021. **G**, COVID-19 TDHOR and HOMR for US states within the 7-month period. **H**, LTC CFR and LTC-to-total ratio for COVID-19 deaths in the US between Jun 26, 202- and Mar 1, 2021. The LTC-total death ratio declined during this outbreak period.

We were interested in whether TDHOR could reflect the trend of HOMR. HOMRs in 34 US states highly correlated with TDHOR, with a 0.99 coefficient of determination (Fig. 1G). The long-term facility (LTC) CFR and LTC-to-total death ratio declined from June 26, 2020 to Mar 1, 2021 (Fig. 1H). This indicated that the prevention in LTC improved in this period. TDHOR and CFR correlated well between 34 states and all US states with coefficients of determination of 0.96 and 1.00, respectively (Appendix p1).

### Early volatility of global CFRs can be attributed to undiagnosed cases

In view of the fact that CFR has large temporal and spatial variations, we were interested in how much variation exists for TDHOR and HOCR. Since CFR = TDHOR* HOCR =TDHR*HCR (formula 10 in Methods 1), there are two contributing factor pairs causing major fluctuations in CFR (TDHOR* HOCR or TDHR*HCR). CFRs for most of these 28 countries can be divided into two stages, a more volatile, or dramatic change stage, followed by a stagnant, or relative flat stage. COVID-19 CFRs in these countries fluctuated considerably over time as the pandemic progressed (Fig. 2A). TDHORs did not exhibit dramatic changes with outbreak stage (Fig 2B).

**Fig. 2.**
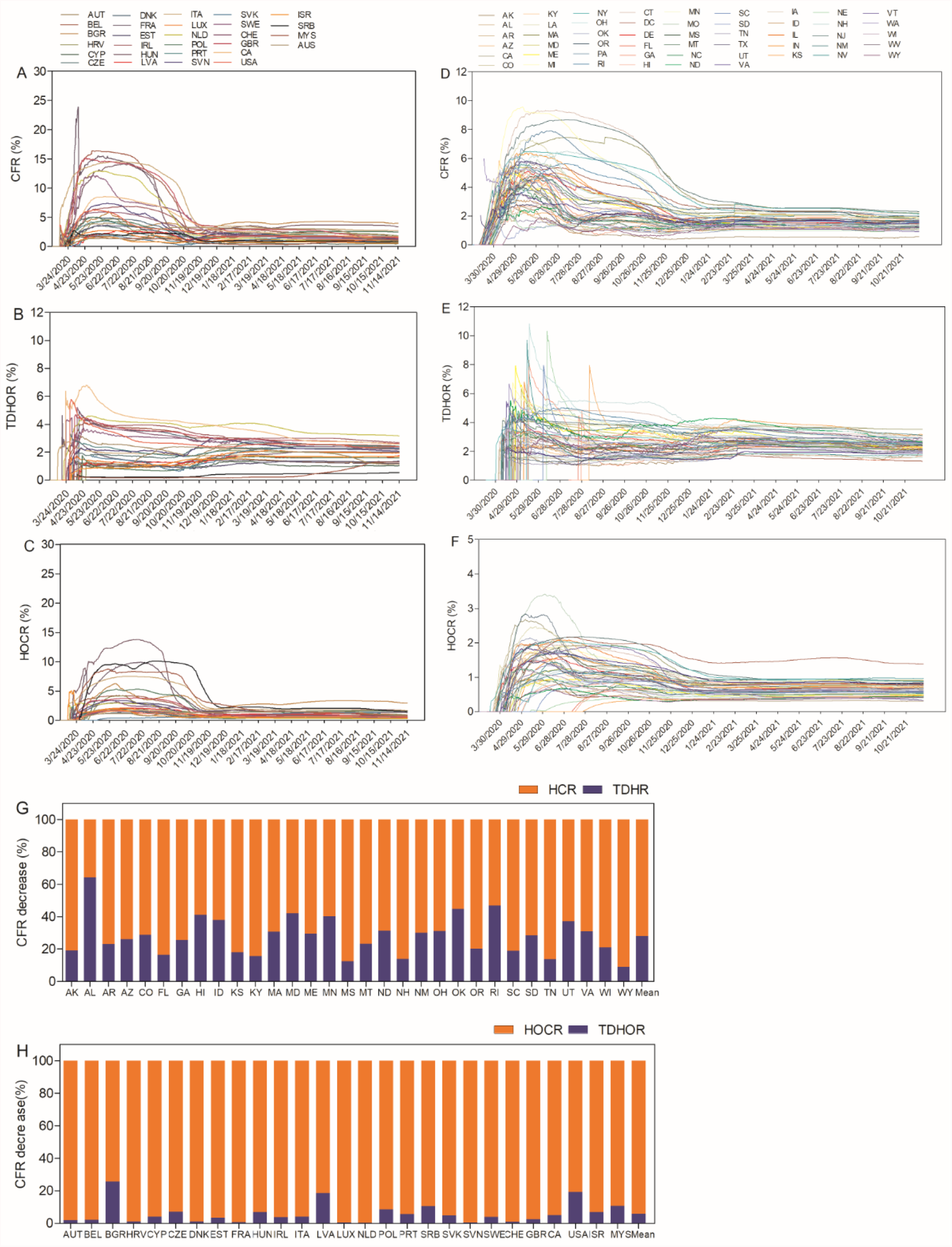
Global and statewide comparisons of COVID-19 CFR, TDHOR, and HOCR. **A**, CFR, **B**, TDHOR, and **C**, HOCR data for COVID-19 among the US, Canada, Israel, Malaysia and 23 European countries between Feb 24, 2020 and Nov 14, 2021. **D-F**, COVID-19 **D**, CFR **E**, TDHOR and **F**, HOCR in all US states/district. **G, H**, Quantification of the CFR decrease due to HCR and TDHR variables for 31 states (including a combined average) between May 2020 and Dec 2020 and 27 countries (between May 2020 and Nov 2020).

Here, we first introduced another new metric, named the “range1-to-mean2 ratio”, to measure the variation of CFR and TDHOR for two stages (dramatic change stage and flat stage, Appendix p2). The average range1-to-mean2 for CFRs of 27 countries was 3.57±1.78 (Methods 4, 5). The average range1-to-mean2 ratio for 27 countries TDHORs was 0.80±0.22, which was smaller than the average range1-to-mean2 of CFR, indicating TDHOR are spatially comparable between countries (Appendix Table 2). HOCR showed a similar pattern to CFR (Fig 2C).

The more consistent TDHOR index, compared to the highly volatile CFR and HOCR metrics, suggest that CFR variations are majorly derived from HOCR. Therefore, we sought to quantitatively analyze the contribution to the dramatic change of CFRs. We examined CFR, HOCR and TDHOR in 27 countries and US states (Fig. 2D-F). The approximate 78.66% decrease in CFR was from HCR (hospitalization-to-case ratio) and 21.34% from TDHR (ratio of total death to hospitalization, Fig. 2G) in USA 31 states.

There are two possibilities for HCR to contribute majorly to the CFR dramatic decrease because HCR equals hospitalization/ (total symptomatic cases-undiagnosed symptomatic case). We hypothesized undiagnosed symptomatic cases maybe the major reason for CFR dramatic decrease, creating an artificially high CFR since they all developed into a low and flat CFR stage after eight to nine months. If the HCR decrease was majorly caused by a decrease in the ratio of hospitalizations within the total symptomatic cases (also means the severe cases rate dramatic decrease), then the LTC CFR would dramatic decrease accordingly, which was not consistent with the LTC CFR (Fig. 1H). There was no new COVID-19 variant nor a dramatic shift in cases to a different age group reported in this period, which could cause the severe cases rate to change dramatically. The change in undiagnosed, symptomatic cases should be the major reason for the dramatic change stage for CFR, confirming our hypothesis.

Similarly, HOCR contributed 89.37% and TDHOR contributed 10.63% to the CFR changes in 27 countries (Fig. 2H, Appendix Table 3). It is reasonable to assume that the major contributing factor for the 27 countries with highest peak CFRs came from undiagnosed, symptomatic case numbers because the TDHORs did not have the same dramatic change during this time.

### COVID-19 fatality monitoring using TDHOR for more than 20 months detected obvious elevations in 17 countries when Alpha variant was reported

The COVID-19 pandemic has progressed as a series of waves, making healthcare facilities overwhelmed and staffing shortages inevitable in certain regions and time periods over the last 20 months. Therefore, fatality risk monitoring and surveillance for short-term and longer-term COVID-19 trends is critical.

To explore the possibility of using TDHOR for COVID-19 fatality risk monitoring, we set a criterion for elevation as values of three continuous days above 30% of the previous three consecutive days from Apr 1, 2020 to Nov 15, 2021. For the TDHOR in the 17 countries that met this elevation criterion, which match the time when Alpha variant was reported (samples taken in September, 2020 in the United Kingdom)^25^, 11 showed no elevations (Fig. 3A). 13 countries had CFR elevations, and 15 showed no CFR elevations (Fig. 3B). Nine countries had HOCR elevations, while 19 countries showed no HOCR elevations (Fig. 3C, Appendix p3,). HOCR and TDHOR provided additional information because they mimic HCR and HMR (Appendix Table 4).

**Figure 3.**
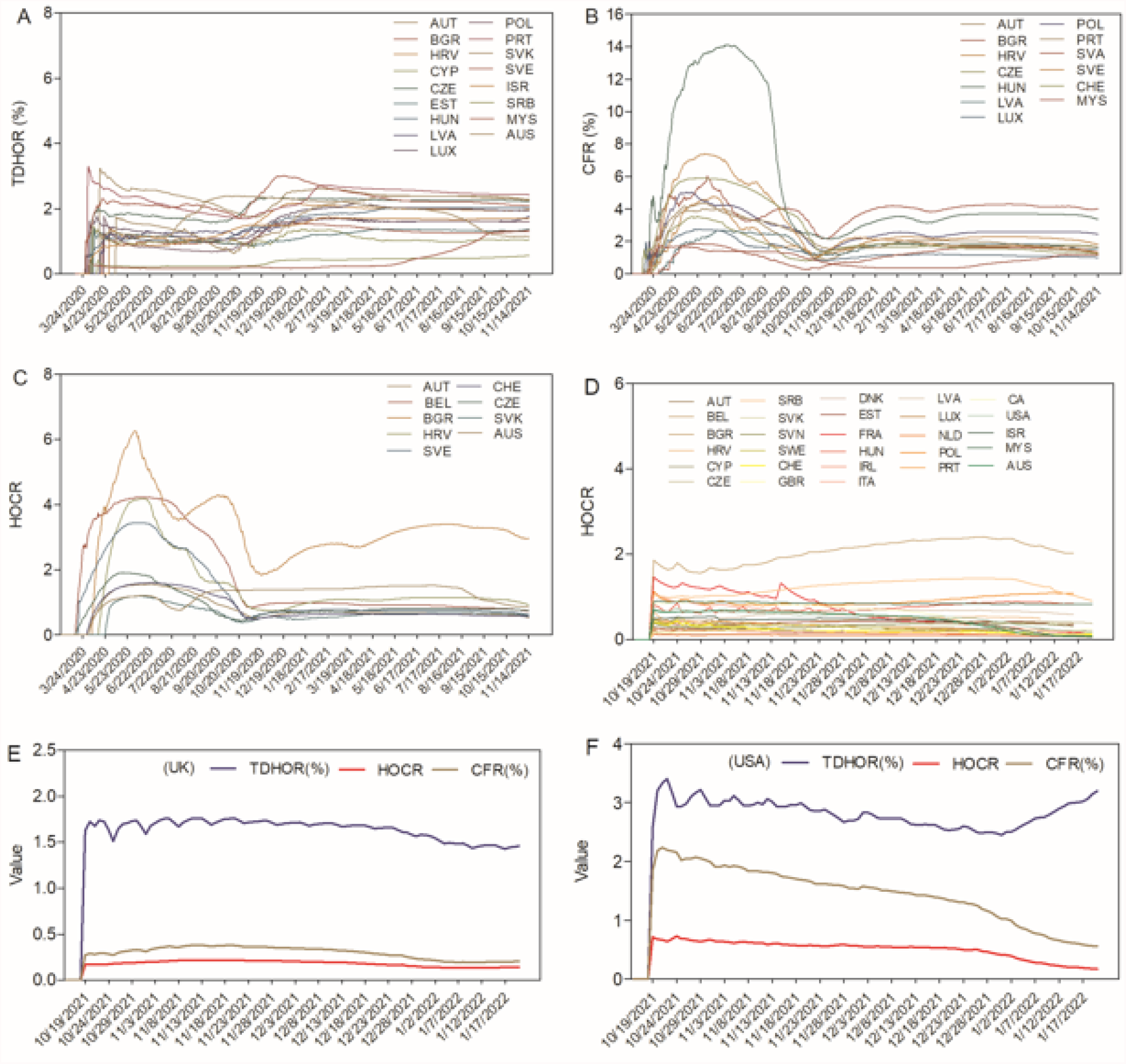
Using TDHOR and HOCR to monitor COVID-19 risk and Omicron variant-associated hospital rate. **A-C**, risk monitoring for 28 countries from Mar 24, 2020 to Nov 15, 2021 with **A**, TDHOR. **B**, CFR, and **C**, HOCR elevations. **D-F**, Comparison of TDHOR, HOCR, and CFR from Oct 16, 2021 to Jan 20, 2022. **D**, HOCR for 28 countries. **E**, TDHOR, HOCR, and CFR for UK. **F**, TDHOR, HOCR, and CFR for USA.

The TDHOR was able to detect elevations for 16 states in the US between Sep 1, 2020 and Mar 1, 2021, while CFR only detected elevations in five states. We could confirm this data for 14 states by HMR and TDHR among 16 states detected TDHOR elevations. The remaining two states (Missouri and Vermont) were unconfirmed since these states did not have cumulative hospitalization data to include in our analysis (Appendix p4). Interestingly, TDHOR and CFR decreases were showed in some US states between Aug 1, 2021 and Nov 15, 2021(all less than 20%) while there was no obvious decrease in HOCR within this period (Fig. 2D-F).

### Monitoring of Omicron variant-associated risk using HOCR showed hospitalization-to-case ratio decreased from Nov 15, 2021 to Jan 16, 2022 in the UK and USA

Genomic surveillance has shown that Omicron variant has spread to the many countries after Omicron was first discovered on Nov 24, 2021 based on samples collected on Nov 11, 2021 in South Africa^15, 26^. It raises serious concerns due to a much higher transmission rate and potential immune escape compared to the Delta variant^27^.

The omicron variant accounted for 58.6% of US cases as of Dec 25, 2021, as estimated by the US CDC ^28^. One of the most important questions is whether the variant increases severe rate or hospitalization-to-case ratio (HCR). The average length of stay in hospital of US (L_mean_) was constant within 7 months (Fig 1E). The L_mean_ of the United Kingdom was also relative steady within 18 months (Appendix p5). These indicate hospitalization-to-case ratio (HCR) can be monitored using HOCR for UK, US and other countries since HOCR equals HCR times *L*_*mean*_.

We measured the TDHORs and HOCRs of the most recent three months for 28 countries from Oct 16, 2021 to Jan 16, 2022 and compared them to CFRs (Fig.3D, Appendix p6). As of Jan 16, 2022, 12 among 28 countries were found to have more than 50% decrease of HOCR compared with the week mean HOCRs of the first week November, 2021(as of Nov 7, 2021). In particular, the UK and the US had decreases in mean week HOCR by 34.08% and 65.16% respectively (Fig.3E-F, Appendix Table 5), which mimics the hospitalization-to-cases ratio in these countris.

## Discussion

It is of the utmost importance to accurately assess and understand the risk for novel diseases like COVID-19, which continues to be a prominent threat to global health. As a novel alternative for COVID-19 risk assessment, we have introduced HOMR, TDHOR and HOCR as three new indexes for COVID-19 fatality risk assessment. TDHOR is valuable as a risk measurement in early stages of COVID-19 outbreaks and has the potential to monitor SARS-CoV-2 mutations that affect the death rate.

Here, in addition to describing the concept and the relationships of HOMR, TDHOR, HOCR, CFR, and HMR, we have applied them to estimate the length of hospital stay in 34 states. The period between Jun 26, 2020 and Aug 6, 2020 did not show a good linear correlation between 1/HOMR and *N*_*dis*_*/ N*_*dead*_. This could reflect either that *f(N*_*hc*_)/*N*_*dead*_ and *N*_*hc*_*/N*_*dead*_ need to be included in the calculation of HOMR or that missing hospitalization information in this data set have a big effect on estimation of *L*_*dis*_ and *L*_*dead*_ during this time period. Together, this is important information for decision makers to properly allocate healthcare resources for COVID-19, as well as for other future infectious diseases.

HOMR showed a high correlation with TDHOR, and imitated HMR well in 34 US states within the first 7 months of the COVID-19 outbreak. This allows us to use TDHOR as another assessment of COVID-19 fatality risk. Based on our analysis, more than two-thirds of the dramatic changes in CFR were caused by undiagnosed cases in USA, while less than one-third were attributed to case-independent data. Notably, TDHORs are less spatially and temporally variable than CFRs on a global level (among 27 countries analyzed), supporting our hypothesis and providing additional value in fatality risk dynamic monitoring. This is additionally supported by CFR data from USA long term care facilities, where COVID-19 cases are far less likely to be undetected, and therefore had no dramatic change stage.

Numerous variants have been identified for the SARS-CoV-2 virus, including the highly transmissible Delta variant ^14^ and the newly-detected Omicron variant^15^. We observed TDHOR elevations in 17 of 28 countries analyzed, along with 16 of the 50 US states in 20 months monitoring. Although there is no direct evidence to link the increase in TDHOR values to the origin of new Alpha variants, this is a possibility that warrants further investigation. The HOCRs of the US and other 11 countries decreased more than 50%, suggesting that Omicron does decrease the hospitalization-to-case ratio as of Jan 16, 2022. This is consistent with other reports about omicron risk^29^.

The criteria for COVID-19 hospitalization in different countries may affect HOMR and TDHOR. The overwhelming healthcare conditions brought about by COVID-19 also restricted available hospitalization capacity at surge times for COVID-19. HOMR, TDHOR and HOCR provide additional information for disease fatality risk assessment and monitoring that are complementary to CFR. It would be interesting to compare HOMR, TDHOR and HOCR for other viruses like seasonal influenza, SARS-CoV-1, MERS-CoV, or Ebola, as well as future, unknown, and potentially fatal diseases. Overall, these new indexes of hospital occupancy related metrics provide the public health sector with additional, effective indicators for monitoring COVID-19 fatality risk, possibly encouraging more countries to release hospital occupancy data in the future since these calculations require data that is relatively easy to collect.

## Data Availability

All data produced in the present study are available upon reasonable request to the authors

## Acknowledgments

The authors would like to thank Dr. Xianglong Kong and Dr. Sheila West for useful discussion and constructive suggestions.

## Author contributions

PWZ initiated ideas and concepts. PWZ, SHZ, WFL, CJK, FT, SL and CAB collected data and did analysis, JS and JW contributed to statistic and epidemiological modelling choices. DJZ and PWZ supervised the whole process of data analysis, results interpretation and manuscript writing.

## Data availability

The authors declare that all data generated or analyzed during this study are included in this published article and its supplementary information files.

## Methods

### 1. Data sources

All data are updated as of Jan 20, 2021, unless specifically mentioned in the text. These data included cases, deaths, current hospital bed occupancy, and cumulative hospitalizations. Symptomatic and asymptomatic cases in mainland China.

Global data, which includes the US but excludes China, was collected from “Our World in Data” (https://ourworldindata.org/coronavirus), which was derived from Johns Hopkins Coronavirus Resource Center and European Centre for Disease Prevention and Control based on its descriptions. Cases, deaths, and hospitalizations for 28 countries analysis were confirmed for COVID-19.

Chinese data was manually collected from the Chinese government by looking at authorized daily announcements for COVID-19 in Chinese areas (National Health Commission of China, http://en.nhc.gov.cn/news.html). Cases, deaths, and hospitalizations for 28 countries analysis were confirmed for COVID-19.

US COVID-19 information was gathered from the “COVID-19 tracking project” before Mar 2, 2021(directly from the websites of US state/territory public health authorities, https://covidtracking.com). After Mar 2, 2021, cases and death data were collected from the US Centers for Disease Control and Prevention, and hospital data were taken from the US Department of Health and Human Services (HHS) (https://healthdata.gov/Hospital/COVID-19-Reported-Patient-Impact-and-Hospital-Capa/g62h-syeh).

### 2. Additional formulas and mathematical derivation for analysis

Formula 3:

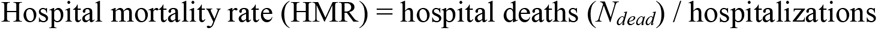

Where:

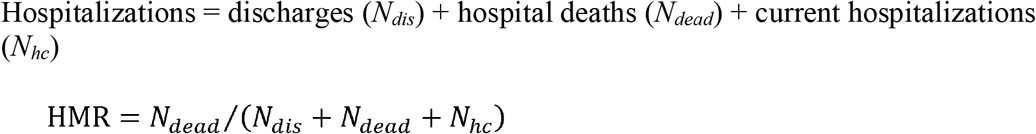

Formula 4:

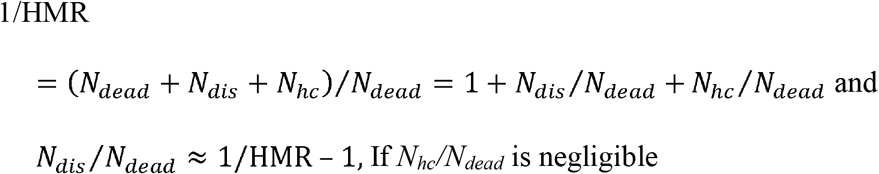

Formula 5:

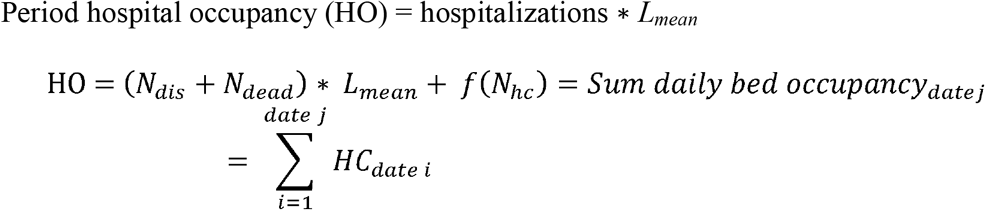

Formula 6:

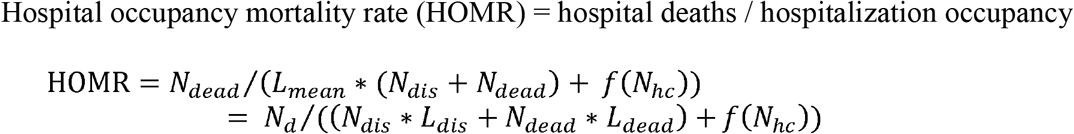

Formula 7:

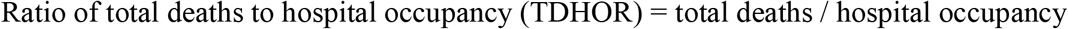

Where:

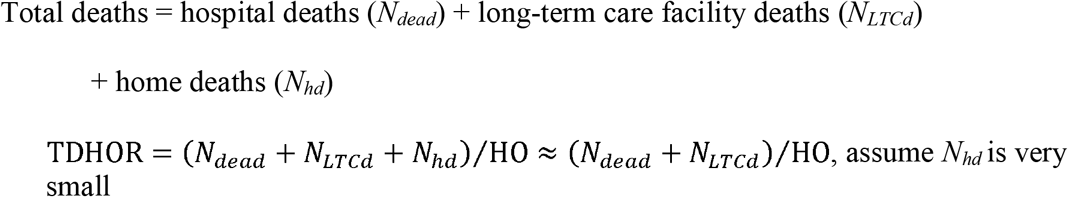

Formula 8:

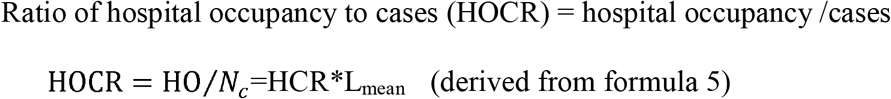

Formula 9:

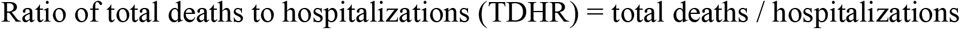

Formula 10:

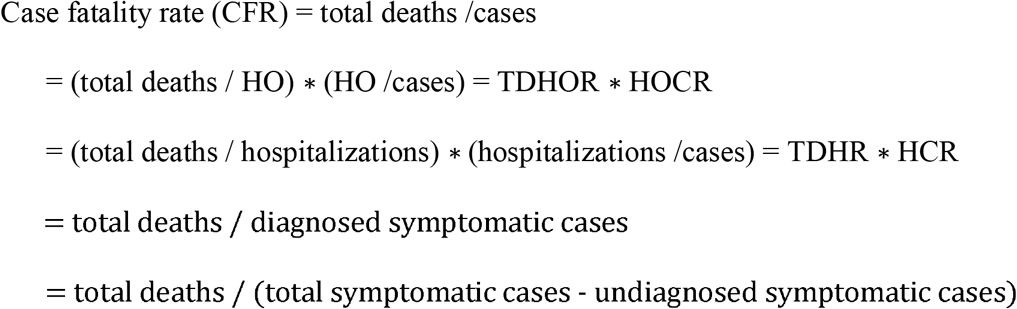

Formula 11:

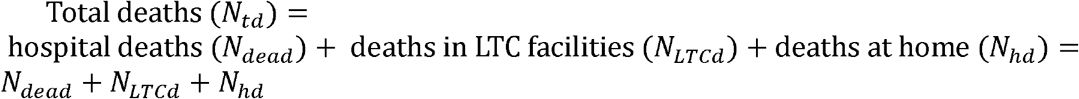

Formula 12:

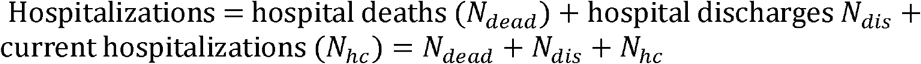

Formula 13:

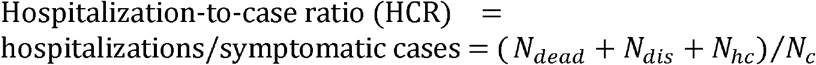

Note: 1) deaths, case and hospitalizations are all cumulative and cases are symptomatic cases if not specified. 2) Long-term care (LTC) facilities includes nursing homes, assisted living, and other long-term care facilities. *N*_*LTCd*_ is the deaths in LTC facilities and *N*_*hd*_ is the deaths at home with COVID-19.

These formulas demonstrate that: 1) HMR is determined majorly by the *N*_*dis*_*/N*_*dead*_ ratio (formula 4) and HOMR depends on *L*_*dead*_, *L*_*dis*_, and the *N*_*dis*_*/N*_*dead*_ ratio and 2) The difference between HOMR (hospital deaths / hospital occupancy) and TDHOR ((hospital deaths + LTC deaths)/ hospital occupancy) is the LTC deaths. HOMR = TDHOR if LTC deaths equals zero. CFR can be segregated by TDHOR * HOCR and TDHR * HCR based on mathematical derivation (formula 10). Under the condition of *N*_*hc*_ is negligible or *N*_*dead*_ >> *N*_*hc*,_ 1/HOMR = *L*_*dis*_ * (*N*_*dis*_/ *N*_*dead*_) + *L*_*dead*_ = *L*_*dis*_ * (1/HMR - 1) + *L*_*dead*_.

### 3. Estimating of average length of stay and comparison with reported data

Formulas 1, 10 and 11 were used for estimation of *L*_*dead*_ and *L*_*dis*_. We assume deaths at home (*N*_*hd*_) for COVID-19 was very low compared to total cases and let total deaths (*N*_*td*_) equal the sum of *N*_*dead*_, *N*_*LTCd*_, and *N*_*td*_ ≈ *N*_*dead*_ + *N*_*LTCd*_ for deaths calculation in our estimation of average length of stay.

Cumulative hospitalization data are needed to calculate HOMR. Only UK and 35 states in the US provide data on cumulative hospitalizations. New Jersey was excluded from our analysis because it had a dramatic change in hospitalizations compared to the other states (New Jersey first released cumulative hospitalization data on May 26, 2020 at 16,373, which jumped from 24517 to 37222 on Oct 22, 2020). The US is the only country with publicly reported LTC deaths. May 21, 2020 was the first day with available LTC data in the US, and Alabama released its first LTC data of 412 deaths on June 25, 2020. The “COVID-19 tracking project” collected LTC data for COVID-19 until Mar 7, 2021; therefore, we used LTC data between Jun 26, 2020 and Mar 1, 2021 for the length of hospital stay estimations. This period covers the highest wave of new COVID-19 cases in the past 20 months. LTC data were smoothed by week before analysis.

First, HO was calculated. Next, *N*_*dead*_ were obtained by subtracting LTC deaths (*N*_*LTCd*_) from total deaths (*N*_*td*_) and calculated HODR using HO and *N*_*dead*_ (where *N*_*dead*_ equals *N*_*td*_ – *N*_*LTCd*_). HMRs were then calculated using cumulative hospitalizations and hospital deaths to get *N*_*dis*_/*N*_*dead*_ using *N*_*dis*_/*N*_*dead*_ ≈ 1/HMR-1(formula 4) and *N*_*dis*_ = cumulative hospitalizations – *N*_*dead*_. Linear regression was performed from Jun 26, 2020 to Mar 1, 2021 using 1/HOMR and *N*_*dis*_/*N*_*dead*_. The plotted trendline was used to determine the average length of stay in hospitals for deaths (*L*_*dead*_) at the intersection of the Y axes. The length of stay in hospitals for discharges (*L*_*dis*_) was determined by the trendline slope. Three different time periods were divided according to these plotting results and their respective r^2^ values.

### 4. Range1-to-mean2 ratio calculation

A new metric, “range1-to-mean2 ratio” (R1/M2), was used to measure the dispersion of CFR and TDHOR for two stages (phases). For this, the range was calculated from the first stage, and the mean was derived from the second stage. Here, we use R1/M2, rather than MR1/M2 (mid-range1 to mean 2 ratio), or Q3R/M2 (Q3 range1 to mean2 ratio) because these calculations are largely similar in this situation.

For this calculation, the range 1 is derived from the first, dramatic change stage (Mar 9, 2020 to October 31, 2020), while the mean2 is calculated from the month immediately following the dramatic change (November, 2020), the second, flat stage (Appendix p2).

### 5. CFR, TDHOR and HOCR variation analysis for 27 countries

The first 14 days of each dataset were omitted if the data was not zero for all analyses due to variations in disease onset, unless indicated in the text. The onset effects of TDHOR were caused by the time gap of when the first daily hospital occupancy data was released and the first death. All CFR, HCR, HOMR, TDHOR, and HMR calculations were cumulative unless specified. HOMR is not available for this analysis due to the lack of LTC data in other countries, except in the US.

Globally, a total of 35 countries have released daily hospital occupancy and deaths data. Eight countries were excluded for analysis. Of these, six European countries, including Finland, Iceland, Lithuania, Norway, Spain, and Malta, had some data missing for either weekends or certain days. Australia does not have a flat stage followed by dramatic change stage in CFR and only had one death in November. It was excluded from analysis. In the case of China, a country that has had a unique COVID-19 progression and time window. It only had four COVID-19 deaths after Apr, 18, 2020.

In total, 27 countries were used for TDHOR, CFR, and HOCR comparison as a single group.

### 6. Early volatility of CFR analysis using TDHOR/HOCR and TDHR/HR

#### 6.1 Cause of CFR volatility analysis using TDHR and HCR for 31 US states

These metrics for all US states and one district showed a similar pattern as the 28 countries previously analyzed, except with some TDHOR onset effects in some states. Based on CFR= HCR*TDHR (formula 10), we first quantitatively analyzed COVID-19 HCR and TDHR contributions in the US. We examined CFRs changes between two months (May, 2020 and December of 2020) in US 35 states for two reasons: 1) only 35 states had cumulative hospitalization data for HR and TDHR calculations and 2) May 2020 was the peak month for CFR and December 2020 was the month after the dramatic change stage for most of the US states.

Four states (Indiana, Nebraska, New Jersey, and Washington) did not have complete cumulative hospitalization data in May 2020. CFR, TDHR, and HCR were calculated for the remaining 31 states for two months (May 2020 and December 2020). Fold decrease for TDHR and HCR between May 2020 and December 2020 were used to calculate the contributions to the CFR changes.

#### 6.2 Cause of CFR volatility analysis using TDHOR and HOCR for 27 countries

From Mar 9, 2020 to Nov 1, 2020, dramatic changes for CFR in 27 countries occurred within an eight-month period, followed by a stagnant stage after Nov 1, 2020. Australia was excluded from analysis because of the same reason mentioned in Methods 5.

Based on CFR= TDHOR*HOCR (Formula 2), we quantitatively analyzed COVID-19 TDHOR and HOCR contributions in these 27 countries and examined CFR changes between two months (May 2020 and November 2020). CFR, TDHR, and HCR were calculated for 31 states for May 2020 and November 2020. Fold decrease for TDHR and HCR between May 2020 and November 2020 were used to calculate the contribution to the CFR changes.

### 7. Fatality risk monitoring using TDHOR

To explore the possibility of using TDHOR and HOCR for COVID-19 fatality risk monitoring, we set the criterion for elevation as values of three continuous days above 30% of the previous three consecutive days among 28 countries between Mar 24, 2020 and Nov.15, 2021.

### 8. Omicron monitoring using HOCR

A criterion was set for elevation or decrease in HOCR as the values of three continuous days after Nov 15, 2021 above or below 30% of the three consecutive days before Dec 31, 2021 for 28 countries or before Jan 11, 2022 for the UK and US.

## Supplementary Material

### Appendix p1

Comparison of US COVID-19 **A**, CFR and **B**, TDHOR. Both CFR and TDHOR in all US states have a high correlation with the 34 states data (Feb 29, 2020 to Mar1, 2021). R^2^ values for each calculation are included in the graphs.

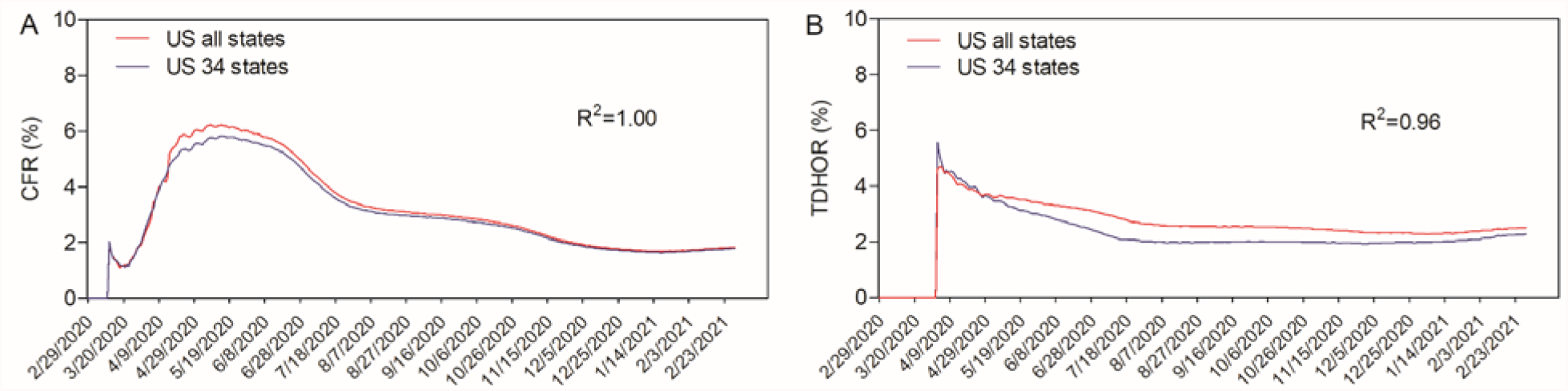

### Appendix p2

Representative **A**, CFR and **B**, TDHOR plots for the COVID-19 outbreak in Hungary. To calculate the R1/M2 ratio, the range was obtained from the initial, volatile outbreak period (stage 1), while the mean was calculated from the second, stagnant phase (stage 2). The start and end points for these phases are denoted by the black lines.

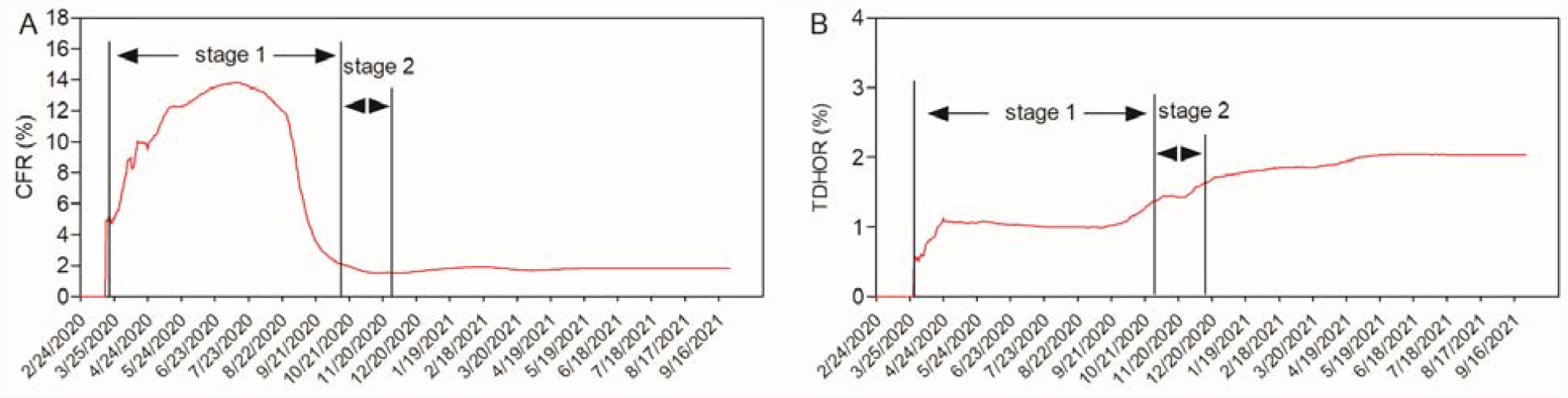

### Appendix p3

**Countries with no elevations for COVID-19 CFR, TDHOR, and HOCR during the outbreak period. A**, No TDHOR elevations for 11 countries **B**, No CFR elevations for 15 countries, and **C**, HOCR in 19 countries with no elevations

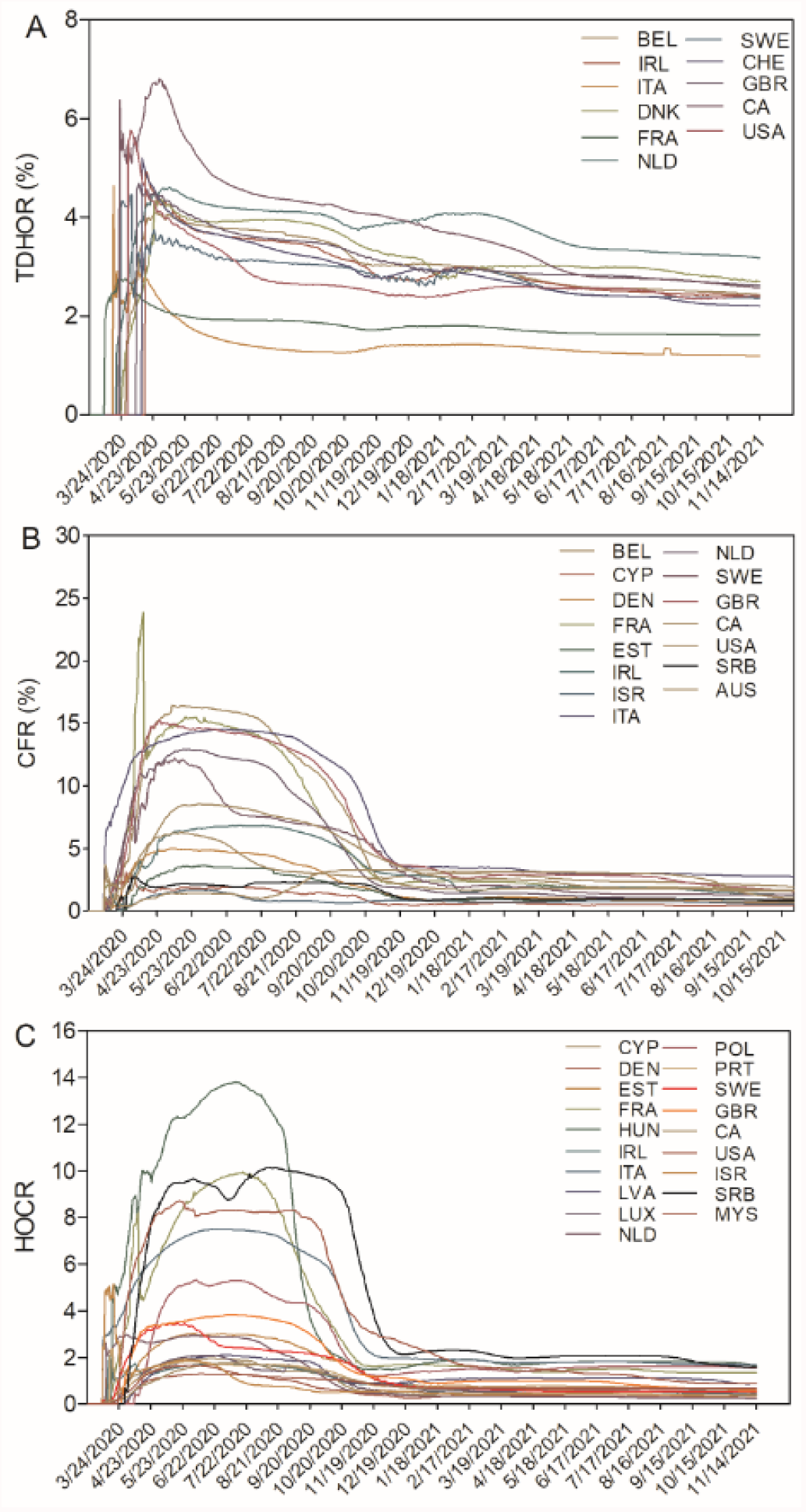

### Appendix p4

**Detection and confirmation of CFR and TDHOR in US states between Sep 1, 2020 and Mar 1, 2021. A**, CFR for 5 states within the US showing elevations during the time course. **B**, COVID-19 TDHOR for 16 US states showing elevations during the same period. **C, D**, 14 states among 16 states with TDHOR elevations were confirmed with TDHR and HMR elevations.

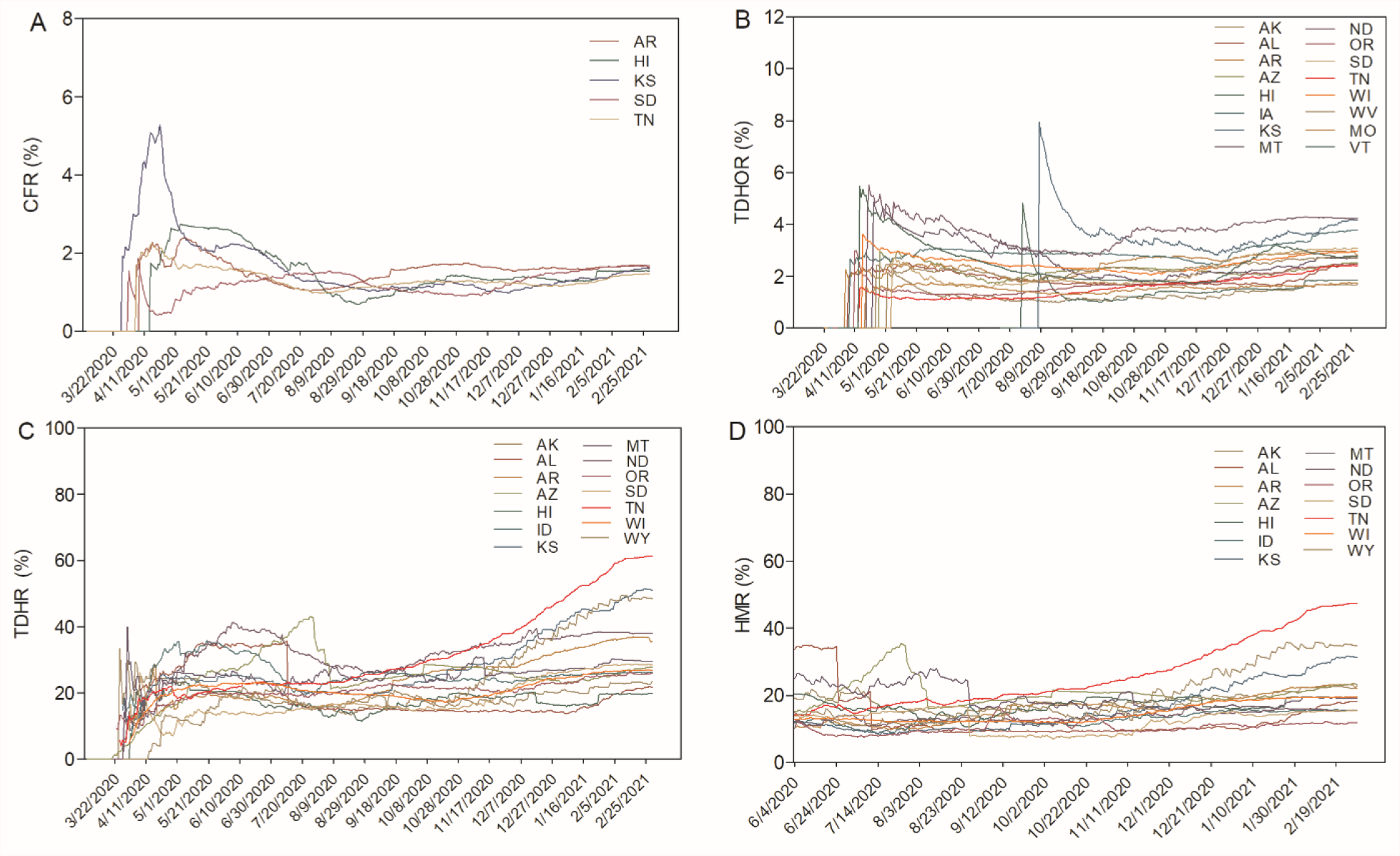

### Appendix p5

**Average length of stay in hospital for COVID-19 deaths and discharges in the United Kingdom from Jun 1, 2020 to Jan 16, 2022**

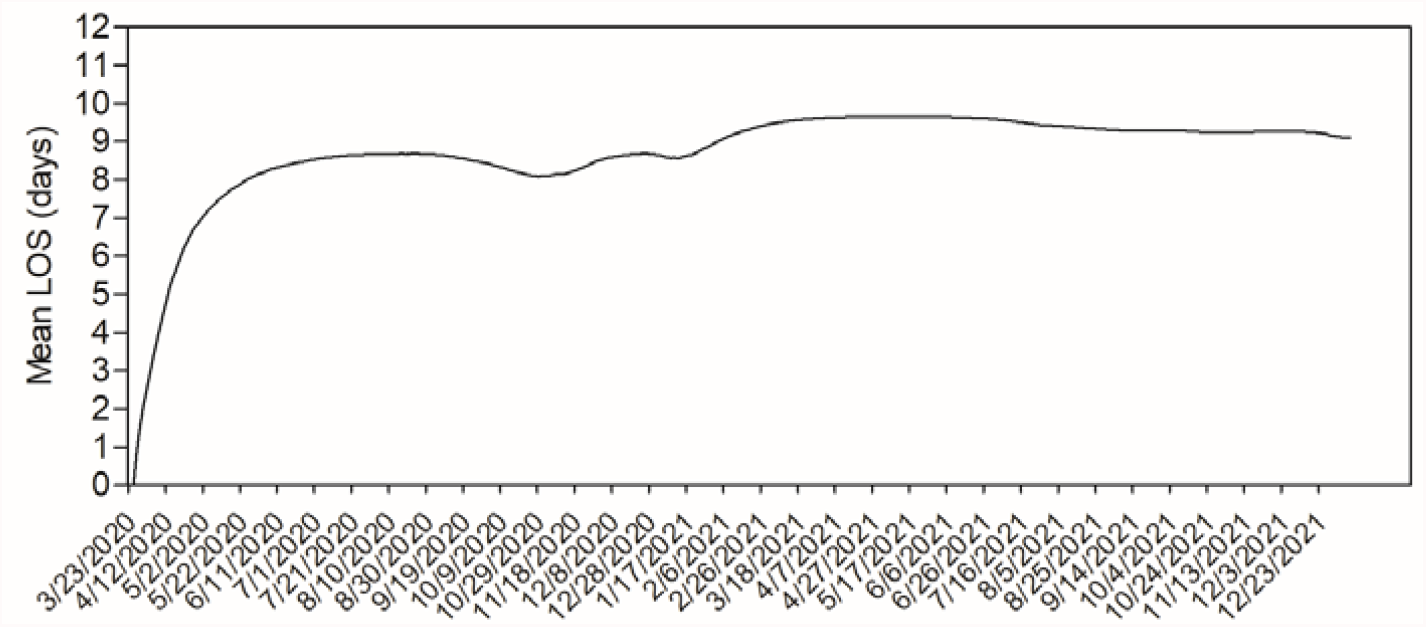

### Appendix p6

**Monitor COVID-19 fatality risk using TDHOR and HOCR from Oct 16, 2021 to Jan 20, 2021. A**, TDHORs and **B**, CFRs for 28 countries.

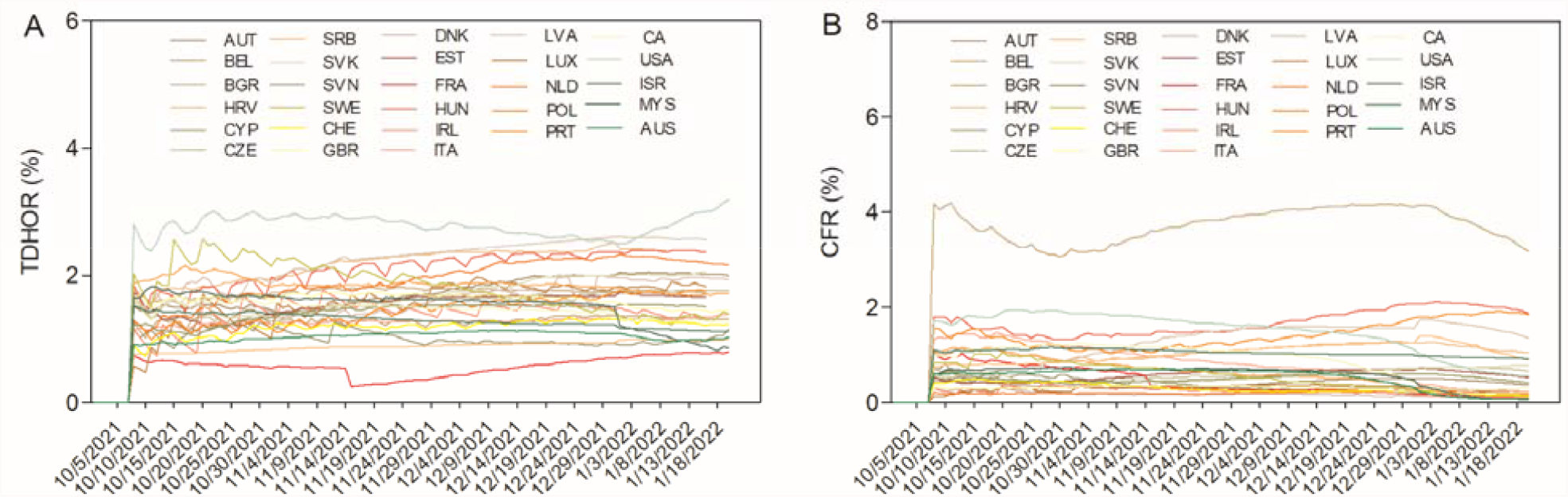

**Appendix Table 1.** Asymptomatic COVID-19 cases in mainland China within a 20-month period.

| Mainland China | Symptomatic cases | Asymptomatic cases | Asymptomatic cases/total cases (%) |
| --- | --- | --- | --- |
| Apr 18, 2020 – Jan 23, 2022 | 22,895 | 14,151 | 38.23% |

**Appendix Table 2.** COVID-19 CFR and TDHOR variations in 27 countries.

|  | CFR |  |  |  | TDHOR |  |  |  |
| --- | --- | --- | --- | --- | --- | --- | --- | --- |
|  | Peak | Range (R1) | November mean (M2) | R1/M2 | Peak | Range (R1) | November Mean (M2) | R1/M2 |
| Austria | 4.02 | 3.9 | 0.97 | 4.03 | 3.22 | 1.49 | 1.81 | 0.82 |
| Belgium | 16.42 | 15.97 | 2.73 | 5.85 | 4.1 | 2.61 | 3.05 | 0.86 |
| Bulgaria | 6.05 | 5.03 | 2.35 | 2.14 | 1.59 | 0.72 | 1.18 | 0.61 |
| Croatia | 4.76 | 4.53 | 1.25 | 3.62 | 2.17 | 1.18 | 1.59 | 0.74 |
| Cyprus | 3.09 | 2.64 | 0.51 | 5.14 | 1.48 | 0.67 | 0.95 | 0.71 |
| Czechia | 3.54 | 3.46 | 1.33 | 2.6 | 2.35 | 0.79 | 2.27 | 0.35 |
| Denmark | 5.02 | 4.9 | 1.22 | 4.0 | 4.94 | 2.23 | 2.9 | 0.77 |
| Estonia | 3.67 | 3.52 | 1.10 | 3.19 | 1.39 | 0.52 | 0.91 | 0.58 |
| France | 23.84 | 22.20 | 2.27 | 9.76 | 2.91 | 1.66 | 1.35 | 1.23 |
| Hungary | 14.11 | 12.74 | 2.21 | 5.78 | 1.74 | 1.23 | 1.43 | 0.86 |
| Ireland | 6.83 | 6.44 | 2.93 | 2.2 | 4.36 | 3.86 | 3.34 | 1.15 |
| Italy | 14.53 | 11.06 | 4.06 | 2.73 | 2.76 | 1.04 | 1.73 | 0.60 |
| Latvia | 2.72 | 2.53 | 1.21 | 2.09 | 1.63 | 1.03 | 1.33 | 0.78 |
| Luxembourg | 2.76 | 2.16 | 0.86 | 2.50 | 1.88 | 0.83 | 1.33 | 0.63 |
| Netherlands | 12.92 | 11.92 | 1.89 | 6.31 | 4.61 | 3.22 | 3.82 | 0.84 |
| Poland | 5.05 | 4.12 | 1.52 | 2.71 | 1.77 | 1.09 | 1.17 | 0.93 |
| Portugal | 4.37 | 4.14 | 1.58 | 2.62 | 3.30 | 1.61 | 1.83 | 0.88 |
| Serbia | 2.71 | 2.49 | 1.22 | 2.04 | 0.42 | 0.22 | 0.27 | 0.79 |
| Slovakia | 1.88 | 1.67 | 0.53 | 3.12 | 1.74 | 1.11 | 1.08 | 1.03 |
| Slovenia | 7.4 | 7.14 | 1.46 | 4.90 | 3.01 | 2.07 | 2.38 | 0.87 |
| Sweden | 12.18 | 12.02 | 3.51 | 3.43 | 4.46 | 2.09 | 2.84 | 0.74 |
| Switzerland | 5.9 | 4.95 | 1.46 | 3.40 | 5.20 | 2.43 | 2.8 | 0.87 |
| United Kingdom | 15.24 | 14.8 | 3.89 | 3.80 | 4.69 | 1.77 | 3.16 | 0.56 |
| Canada | 8.56 | 8.23 | 3.7 | 2.22 | 6.81 | 3.00 | 4.06 | 0.74 |
| United States of America | 6.25 | 4.54 | 2.23 | 2.04 | 5.77 | 3.39 | 2.50 | 1.36 |
| Israel | 1.71 | 1.63 | 0.84 | 1.93 | 1.51 | 1.08 | 1.50 | 0.72 |
| Malaysia | 1.74 | 1.52 | 0.66 | 2.31 | 0.28 |  |  |  |
| Mean±SD | 7.31±5.50 | 6.68±5.15 | 1.83±1.03 | 3.57±1.78 | 2.97±1.68 | 1.60±0.98 | 1.95±1.02 | 0.81±0.22 |

**Appendix Table 3.** ISO ALPHA-3 codes for 28 countries.

| **Countries** | **ISO ALPHA-3 code** |
| --- | --- |
| Australia | AUS |
| Austria | AUT |
| Belgium | BEL |
| Bulgaria | BGR |
| Canada | CAN |
| Croatia | HRV |
| Cyprus | CYP |
| Czechia | CZE |
| Denmark | DNK |
| Estonia | EST |
| France | FRA |
| Hungary | HUN |
| Ireland | IRL |
| Israel | ISR |
| Italy | ITA |
| Latvia | LVA |
| Luxembourg | LUX |
| Malaysia | MYS |
| Netherlands | NLD |
| Poland | POL |
| Portugal | PRT |
| Serbia | SRB |
| Slovakia | SVK |
| Slovenia | SVN |
| Sweden | SWE |
| Switzerland | CHE |
| United Kingdom | GBR |
| United States | USA |

**Appendix Table 4.** List of countries with elevations detected by the different monitoring metrics.

| CFR elevation<br>(13 Countries) | HOCR<br>elevation<br>(9 Countries) | TDHOR<br>elevation<br>(17 Countries) | HOCR or<br>TDHOR<br>elevation (19) | HOCR<br>elevation with<br>no TDHOR<br>elevation<br>(2 countries) | TDHOR<br>elevation with<br>no HOCR<br>elevation<br>(10 countries) | HOCR and<br>TDHOR<br>elevations<br>(7 countries) |
| --- | --- | --- | --- | --- | --- | --- |
| AUT | AUT | AUT | AUT | BEL | CYP | AUT |
| BGR | BEL | BGR | BEL | CHE | EST | BGR |
| HRV | BGR | HRV | BGR |  | HUN | HRV |
| CZE | HRV | CYP | HRV |  | LVA | CZE |
| HUN | CZE | CZE | CYP |  | LUX | SVK |
| LVA | SVK | EST | CZE |  | POL | SVN |
| LUX | SVN | HUN | EST |  | PRT | AUS |
| POL | CHE | LVA | HUN |  | SRB |  |
| PRT | AUS | LUX | LVA |  | ISR |  |
| SVK |  | POL | LUX |  | MYS |  |
| SVN |  | PRT | POL |  |  |  |
| CHE |  | SRB | PRT |  |  |  |
| AUS |  | SVK | SRB |  |  |  |
|  |  | SVN | SVK |  |  |  |
|  |  | ISR | SVN |  |  |  |
|  |  | MYS | ISR |  |  |  |
|  |  | AUS | CHE |  |  |  |
|  |  |  | MYS |  |  |  |
|  |  |  | AUS |  |  |  |

**Appendix Table 5.** Omicron risk monitoring using HOCR.

| **Countries** | **Mean Week**<br>**HOCR as of**<br>**Nov 7, 2021** | **Mean Week**<br>**HOCR as of**<br>**Jan 16, 2022** | **HOCR week Change**<br>**(%, Nov 7, 2021 vs Jan**<br>**16, 2022)** |
| --- | --- | --- | --- |
| **Austria** | 0.23 | 0.22 | -5.51 |
| **Belgium** | 0.24 | 0.20 | -16.45 |
| **Bulgaria** | 1.67 | 2.07 | 24.31 |
| **Croatia** | 0.41 | ND* | ND* |
| **Cyprus** | 0.39 | 0.12 | -69.96 |
| **Czechia** | 0.31 | 0.42 | 31.12 |
| **Denmark** | 0.12 | 0.06 | -54.15 |
| **Estonia** | 0.35 | 0.37 | 4.91 |
| **France** | 1.16 | 0.18 | -84.72 |
| **Hungary** | 0.68 | 0.85 | 25.15 |
| **Ireland** | 0.20 | 0.08 | -60.04 |
| **Italy** | 0.78 | 0.19 | -75.48 |
| **Latvia** | 0.52 | 0.61 | 19.04 |
| **Luxembourg** | 0.21 | 0.12 | -44.35 |
| **Netherlands** | 0.11 | 0.09 | -15.27 |
| **Poland** | 0.85 | 1.08 | 27.79 |
| **Portugal** | 0.41 | 0.10 | -76.13 |
| **Serbia** | 1.04 | 1.13 | 8.51 |
| **Slovakia** | 0.29 | 0.37 | 29.78 |
| **Slovenia** | 0.26 | 0.33 | 27.01 |
| **Sweden** | 0.34 | 0.12 | -66.19 |
| **Switzerland** | 0.29 | 0.13 | -53.51 |
| **United Kingdom** | 0.20 | 0.13 | -34.08 |
| **Canada** | 0.88 | 0.21 | -75.58 |
| **United States** | 0.64 | 0.22 | -65.16 |
| **Israel** | 0.47 | 0.09 | -80.14 |
| **Malaysia** | 0.88 | 0.83 | -5.56 |
| **Australia** | 0.68 | 0.08 | -88.00 |

\*ND, No data.

## Supplementary files

**Supplementary file 1. (separate file)**

Excel file containing 28 countries data for COVID-19(Feb 24, 2020-Jan 20, 2021).

**Supplementary file 2. (separate file)**

Excel file containing COVID-19 data for US states (Feb 24, 2020-Nov 15, 2021)

**Supplementary file 3. (separate file)**

Excel file containing mainland Chinese data for COVID-19(Jan 9, 2020-Jan 23, 2021). A website with mainland Chinese COVID-19 data (including Wuhan and Hubei) is ready to activate.

